# Protocol of a randomized controlled trial testing inhaled Nitric Oxide in mechanically ventilated patients with severe acute respiratory syndrome in COVID-19 (SARS-CoV-2)

**DOI:** 10.1101/2020.03.09.20033530

**Authors:** Chong Lei, Binxiao Su, Hailong Dong, Andrea Bellavia, Raffaele Di Fenza, Bijan Safaee Fakhr, Stefano Gianni, Luigi Giuseppe Grassi, Robert Kacmarek, Caio Cesar Araujo Morais, Riccardo Pinciroli, Emanuele Vassena, Lorenzo Berra

## Abstract

**Introduction:** Severe acute respiratory syndrome due to novel Coronavirus (SARS-CoV-2) related infection (COVID-19) is characterized by severe ventilation perfusion mismatch leading to refractory hypoxemia. To date, there is no specific treatment available for COVID-19. Nitric oxide is a selective pulmonary vasodilator gas used as a rescue therapy in refractory hypoxemia due to acute respiratory distress syndrome (ARDS). In has also shown *invitro* and clinical evidence that inhaled nitric oxide gas (iNO) has antiviral activity against other strains of coronavirus. The primary aim of this study is to determine whether inhaled NO improves oxygenation in patients with hypoxic COVID-19. This is a multicenter randomized controlled trial with 1:1 individual allocation. Patients will be blinded to the treatment.

**Methods and analysis:** Intubated patients admitted to the intensive care unit with confirmed SARS-CoV-2 infection and severe hypoxemia will be randomized to receive inhalation of NO (treatment group) or not (control group). Treatment will be stopped when patients are free from hypoxemia for more than 24 hours. The primary outcome evaluates levels of oxygenation between the two groups at 48 hours. Secondary outcomes include rate of survival rate at 28 and 90 days in the two groups, time to resolution of severe hypoxemia, time to achieve negativity of SARS-CoV-2 RT-PCR tests.

**Ethics and dissemination:** The study protocol has been approved by the Investigational Review Board of Xijing Hospital (Xi’an, China) and by the Partners Human Research Committee (Boston, USA). Recruitment will start after approval of both IRBs and local IRBs at other enrolling centers. Results of this study will be published in scientific journals, presented at scientific meetings, reported through flyers and posters, and published on related website or media in combating against this widespread contagious disease.

**Trial registration:** Clinicaltrials.gov. NCT04306393

**Strengths and limitations of this study:**

— Supplementation with nitric oxide (NO) might improve oxygenation and survival of SARS-CoV-2-infected patients.
— The antiviral activity of NO inhalation will be explored by measuring the time difference between the two groups to reach SARS-CoV-2 rt-PCR negativity.
— The spread of the disease worldwide determines the geographic areas of study and the recruitment rate of patients.

## Introduction

The first pneumonia case with unknown etiology were identified in Wuhan, China in early December 2019 [1]. The genomic sequence of this new pathogen and a diagnostic real-time reverse transcriptase polymerase (rt-PCR) chain assay were published in January 2020. High throughput sequencing of the bronchoalveolar samples from these patients has revealed a novel betacoronavirus that is currently named SARS-CoV-2, as it resembled severe acute respiratory syndrome coronavirus [2].

SARS-CoV-2 has so far infected more than 110,000 people worldwide with more than 4,000 deaths (3.6% of diagnosed patients). The World Health Organization has declared the SARS-CoV-2 infection as a public health emergency of international concern. [3]. The majority of the cases has been reported in China, but significant outbreaks are taking place in South Korea, Japan, Europe, Iran and more recently in USA [4]. In the human host, SARS-CoV-2 infection may be asymptomatic or may cause a syndrome (named COVID-19) ranging from common cold to a severe pneumonia with acute respiratory syndrome and need of mechanical ventilation in intensive care unit (ICU). In a retrospective Chinese study on 138 consecutive patients admitted with COVID-19, the median time from clinical onset to hospital admission was 7 days, 26% of patients were admitted to the ICU and 61% of them met clinical criteria for acute respiratory distress syndrome (ARDS) [5]. Another retrospective Chinese study on critically ill patients with COVID-19 pneumonia showed that 67% of patients met ARDS criteria, with a mortality of 61.5% at 28 days. Reported casualties in the ICU are characterized of various profiles of multiorgan failure (81% of deceased patients had ARDS, 37.5% had AKI, 28% had cardiac injury and 28% had liver failure) [6]. Autoptic findings in a published clinical case showed features resembling those of coronavirus-related infections such as Severe Acute Respiratory Distress Syndrome (SARS) and Middle Eastern Respiratory Syndrome (MERS), including bilateral diffuse alveolar damage with fibromyxoid exudates, desquamation of pneumocytes and hyaline membrane formation. Findings in cardiac and hepatic tissues may suggest the contribution of a viral infection as well [7].

COVID-19 is highly contagious and responsible for thousands of casualties and is now spreading to many countries. The combined effect of the high transmission and the reported high incidence of severe disease in symptomatic patients poses a threat to healthcare systems involved in the outbreaks in different ways, including: cumulative casualties, increased need for hospital and ICU beds causing work overload for all the healthcare staff and often forcing hospitals to shut down all elective surgical activity; high social and economic costs, dramatically reduction of the productivity by the rigorous quarantine requirements and strategies for curtailing disease spreading, which made economic pain that disrupt supply chains and stock markets. To-date, no treatment showed to increase survival and decrease the need for ventilatory support in patients with severe acute respiratory syndrome due to COVID-19. In responding to this epidemic, the treatment strategies should be safe, can be quickly pass through the regulatory reviews and can be used on a massive scale with low cost. Therefore, screen for the existing treatments seemed to be the potential choice.

In clinical settings, NO gas has been approved by the US Federal Drug Administration for the treatment of pulmonary hypertension of the newborn in presence of hypoxic respiratory failure. However, NO gas has been advocated as rescue treatment in adults with hypoxic ARDS [8]. In 2004, during the Severe Acute Respiratory Syndrome Coronavirus (SARS-CoV) outbreak, Chen et al. reported the use of inhaled NO gas (iNO) in six patients with severe symptoms. Treatment with iNO reversed pulmonary hypertension, improved remarkably severe hypoxemia and shortened the length of ventilatory support as compared to matched control patients with SARS-CoV [9]. In a subsequent *in-vitro* study, nitric oxide (NO) donors (e.g. S-nitroso-N-acetylpenicillamine) greatly increased the survival rate of SARS-CoV-infected eukaryotic cells, suggesting direct antiviral effects of NO [9]. Coronavirus responsible for SARS-CoV shares most of the genome of COVID-19 virus indicating potential effectiveness of iNO therapy in these patients.

Due to similarities with the Coronavirus responsible for SARS and COVID-19, we hypothesize that in addition to improve the oxygenation of the severe cases, iNO gas retains potent antiviral activity against SARS-CoV-2 responsible for COVID-19. We designed this study to assess whether continuous delivery of iNO as rescue therapy may increase oxygenation and improves survival in patients with COVID-19.

## Methods and Analysis

### Study Setting

This is a multicenter, randomized (1:1) controlled, parallel-arm clinical trial. Given the absence of a targeted therapy, the refractory hypoxemia and the high mortality associated with SARS-CoV-2, we propose this protocol to all interested centers with SARS-CoV-2 patients.

### Eligibility criteria

Inclusion criteria are: (1) Adult patients, ≥ 18 years old; (2) admitted to the ICU; (3) intubated and mechanically ventilated; (4) confirmed diagnosis by positive rt-PCR of SARS-CoV-2 infection. Exclusion criteria are: (1) Patients intubated for more than 72 hours from initiation of the treatment gas; (2) Subjects enrolled in another interventional research study; (3) Physician of record opposed to enrolling the patient due to perceived safety concerns; or any condition that does not allow the protocol to be followed safely; (4) Subjects with past medical history of lung malignancy or pneumonectomy or lung transplant; (5) Subjects receiving a tidal volume < 3 cc/kg of ideal body weight at the time of enrollment; (6) Subjects with severe burns involving more than 40% of Total Body Surface Area; (7) Subjects that have experienced cardiac arrest with CPR for longer than 30 minutes; (8) Subjects with a presumed severe deficit in cerebral function with fixed dilated pupil; (9) Subjects receiving renal replacement therapy at the time of enrollment; (10) Subjects who have an impaired ability to ventilate without assistance; (11) Subjects who have a history of malignancy or other irreversible disease/conditions with a 6-month mortality > 50%; (12) Subjects not fully committed to full support at the time of enrollment.

### Interventions

Eligible patients are randomized to receive either institutional standard of care with iNO addition (treatment arm) or plain institutional standard of care (control arm). Standards of care are delivered according to the institution own protocols (such as ventilation strategies and use and dosage of antivirals and antimicrobials, steroids, inotropic-vasopressor agents and initiation of extracorporeal membrane oxygenator [ECMO]).

Patients in the treatment arm receive iNO at 80 ppm for the first 48 hours after enrollment, then reduce to 40 ppm until severe hypoxemia resolves. Weaning from NO will start when patients improve the level of oxygenation to PaO_2_/FiO_2_> 300 mmHg for more than 24 hours consecutively. Since abrupt discontinuation of iNO can sometimes result in rebound pulmonary hypertension, with possible oxygenation impairment and acute right heart failure, gradual discontinuation will be performed. Physicians will follow their own institution weaning protocols. In absence of institutional protocols, iNO will be reduced every 4 hours in stepwise fashion starting from 40 ppm to 20, 10, 5, 3, 2 and 1 ppm. If case of hypoxemia (SpO_2_< 93%) or acute hypotension (systolic blood pressure < 90 mmHg) during weaning, iNO should be increased to the prior higher concentration. Weaning can be started earlier, but always according to the institutional protocol or the protocol proposed above, if one of the two following scenarios applies. (1) Fast weaning after the first 48 hours of gas administration can be started if the clinical team foresees an extubation happening within no more than 48 hours, a PaO_2_/FiO_2_> 200 mmHg for more than 24 hours and hemodynamics do not require infusion of more than 10 micrograms of norepinephrine or 2 inotropic/vasopressors to achieve a systolic blood pressure > 100 mmHg and mean arterial pressure > 60 mmHg. (2) Extra-fast weaning within the first 48 hours of gas administration can be started if the ICU team foresees that a newly intubated patient will need to be extubated within the next 12 hours, the patient is on spontaneous ventilation with PaO2/FiO2>300 mmHg, PEEP<10 cm H2O and FiO2<50%, and hemodynamics do not require infusion of more than 10 micrograms/min of norepinephrine or 2 inotropic/vasopressors to achieve a systolic blood pressure > 100 mmHg and mean arterial pressure > 60 mmHg.

After iNO discontinuation, the gas shall be restarted in case one of the following conditions applies: P/F < 200 mmHg; requirement for paralysis; requirement for pronation. Inhaled NO should re-initiate at 40 ppm and weaned according to weaning protocols

### Safety

NO reacts with oxygen to form NO_2_, which may cause airway inflammation and damage to lung tissues. Moreover, NO oxidizes ferrous Hb to form Met-Hb, which is unable to transport and release oxygen to tissues. [11]. The binding of NO to Hb is a rapidly reversible reaction, with a half-life of 15–20 min after NO discontinuation. The side effects and adverse events related to iNO delivery are well reported in the literature. Based on the present literature and Food and Drug Administration reports, the risks of breathing iNO at 80 ppm for 24 hours are minimal when Met-Hb levels and NO/NO_2_ delivery levels are carefully monitored [12]. To improve safety, in the present trial, iNO is administered and monitored by trained clinicians. The NO_2_ will be monitored and maintained at levels of below 2 ppm. Met-Hb is continuously monitored by non-invasive co-oximetry. If Met-Hb levels exceed 5% of circulating Hb, the concentration of NO delivered is halved and closely monitored until a reduction occurs. If MetHb levels persist above 5%, iNO is progressively halved until a reduction below 5% occurs.

### Blinding

The study will be blinded to the patient. Outcome assessors, data analysts, the study personnel delivering the NO gas and monitoring patient’s methemoglobin levels will not be blinded.

### Outcomes

The primary outcome is the improvement of arterial oxygenation at 48 hours versus baseline from enrollment between the two groups. If a patient dies before the initiation of the gas, the patient will not enter the trial. If a patient dies during the first 48 hours of treatment, the last available blood gas analysis will be used. Levels of oxygenation will be calculated by the PaO_2_/FiO_2_ ratio. Patients will be followed until 90 days from enrollment.

Secondary outcomes include: (1) time to reach normoxemia defined by a PaO_2_/FiO_2_ ≥ 300 for at least 24 hours during the first 28 days after enrollment. If a patient dies before day 28, the patient will be considered as “never recovered”; (2) proportion of SARS-CoV-2 free patients (i.e., normoxiemic patients) during the first 28 days after enrollment. If a patient dies before day 28, the patient will be considered as “never recovered”; (3) Survival at 28 days and 90 days from enrollment.

The following data will be collected to assess exploratory outcomes, such as: (1) Daily oxygenation in the two groups until day 28; (2) Need for new renal replacement therapy during the first 28 days; (3) Mechanical support of circulation (i.e., ECMO, intra-aortic balloon pump, VADs) during the first 28 days; (4) Days free of vasopressors during the first 28 days; (5) Ventilator-free day at 28 days; (6) Time to SARS-CoV-2 rt-PCR negative in upper respiratory tract specimen (assessed within the first 28 days); (7) ICU-free days at 28 days.

### Sample size

We hypothesize that NO gas therapy leads to an improvement of oxygenation due to amelioration of ventilation perfusion matching in patients affected with SARS-Co-V2. A difference of 20% in PaO_2_/FiO_2_ ratio between the two groups at 48 hours from enrollment is considered to be clinically relevant. Available data on oxygenation in SARS-CoV and SARS-CoV-2 patients in ICU are limited [5] [6] [13]. A previous study in ARDS patients ventilated according to standard of treatment (ARDSnet table) reported a PaO_2_/FiO_2_ ratio at 72 hours of 190 + 71 mmHg [14]. We hypothesize that iNO gas may increase the PaO_2_/FiO_2_ ratio by at least 20% in the treatment group. Assuming an alpha of 0.05 and a beta (power) of 0.9, we calculated with a two-sample means test (Satterthwaite’s t test assuming unequal variances) a need for N1=91 and N2=91 patients. A 10% of dropouts after ICU discharge is foreseen and sample size is thereby increased to N=200 patients. Estimated sample sizes where calculated using Stata 14.1 software.

### Recruitment

All patients admitted into recruiting ICUs with SARS-CoV-2 and intubated for hypoxemia are screened for eligibility. If a patient is excluded, the reasons are noted on a screening log. To obtain informed consent, the details of the study are presented to the patient’s healthcare proxy. A copy of the consent form will be given to the patient’s healthcare proxy. Prior to the initiation of any study procedures, the proxy’s written consent is obtained by a clinician.

### Assignment of interventions

The randomization sequence is created by an independent statistician using Stata 14.1. A predetermined block randomization method (fixed block size of 10) is used to ensure equal distribution of participants to treatment arms and will be uploaded on REDCap.

### Data collection methods

Clinical information including medical history and laboratory exams will be obtained from the medical charts. Collection of study variables will be managed by the outcome assessors by using a dedicated patient’s file on REDCap.

### Data management

Outcome assessors, treatment providers and the principal investigator will obtain unique usernames and password to transfer all data to a REDCap page dedicated to the study.

### Statistical methods

Data analysis will be based on the Intention-to-treat principle. For patients dying during the first 48 hours of treatment, the last available blood gas analysis will be used to assess the primary outcome. For the proposed trial, we anticipate that no participants will miss the primary evaluation.

Demographic and clinical characteristics will be presented as number (percentage), mean (SD), or median (interquartile range [IQR]). Comparisons between groups will be made using the χ2 test or Fisher’s exact test for categorical variables and T-test or Wilcoxon’s rank-sum test for continuous variables as appropriate. Effect sizes will be described with the probability of more favorable outcome (probabilistic index) and 95% CI will be calculated.

The primary endpoint will be compared using T-test or Mann-Whitney U as appropriate. Time to reach normoxemia will be compared within the two groups with a T-Test or Mann-Whitney U test, as appropriate. To exclude or confirm that the expected benefit in oxygenation by NO may be evident during the first days of treatment and then decrease we will consider the proportion of SARS-CoV-2 free patients during 28 days and compare the two groups in terms of treatment success with a log-rank test. Survival curves will be generated via the Kaplan-Meier method and compared with a log-rank test.

Rates of organ dysfunction will be compared using Fisher’s exact test. Ventilator-free days, ICU-free days, days free of vasopressors and time to negative SARS-CoV-2 rt-PCR will be compared using the T-test or Mann-Whitney U test, as appropriate. Fisher’s exact test will be used to estimate treatment differences in the incidence of each specified adverse event. No adjustments will be made for multiple hypothesis evaluations of safety endpoints. For all analyses, a 2-sided alpha threshold of .05 will be considered significant.

To adjust for additional influent factors not involved in the randomization process such as age, ARDS severity and co-administration of other experimental treatments, we will perform a multivariate logistic regression analysis on the primary outcome.

## Data monitoring

Data quality and safety will be monitored by each site’s PI, the study Medical Coordinating Center (MCC) and a Data Safety Monitoring Board (DSMB). The site PIs will continuously monitor the data quality according to data quality measures. In addition, recruitment rate, deviation from inclusion/exclusion criteria and protocol, and confidentiality of data and database will be monitored. A DSMB, drawing from persons with expertise in the area will be constituted.

An interim analysis will be performed at 50% of planned enrollment. The DSMB will evaluate survival rate between the two groups. TheD SMB will determine whether the study should continue or terminate. The DSMB will play a valuable role in advising the study leadership on the relevance of advances in the diagnosis and treatment of patients. A number of therapeutic or diagnostic testing advances may possibly occur during the course of the trial. If protocol modifications are warranted, close consultation among the DSMB and the study leadership will be required. A separate DSMB charter that outlines in detail the operating guidelines for the committee and the protocol for evaluation of data will be developed prior to the start of patient randomization and agreed upon in the initial meeting of the DSMB.

### Harms

Organ failure, prolonged hospitalization and mortality will be recorded as part of the study defined outcomes. Other adverse events will be monitored by the site PI and research specialist in real time from the start of randomization to hospital discharge or death. Reportable adverse events will include hemodynamic change and deterioration of oxygenation related to NO inhalation and require the adjustment of NO concentration. A serious adverse event in this study will be defined as a reportable adverse event that is fatal, life-threatening, or permanently disabling. All adverse events will be indicated on the data forms for the study and on the specific adverse event report forms and all serious adverse events will be reported to the site IRB within 48 hours of the research team learning about the event followed by more detailed written report to the local IRB. The following information about adverse events will be collected: 1) the onset and resolution of the event, 2) an assessment of the severity or intensity of the event, 3) an assessment of the relationship of the event to the intervention, and 4) any action taken because of event. Sites will inform the principle investigators (CL and LB) at the coordinating centers (Xijing Hospital and Massachusetts General Hospital) for any related adverse events directly caused by inhalation of NO (non-serious) within 14 days of discovery. All fatal, life-threatening, or permanently disabling (regardless of relatedness) events due to inhalation of NO must be reported to the coordinating centers within 48 hours of discovery. The principle investigators will report these events to the DSMB and to the coordinating centers IRBs.

### Patient and Public Involvement

Participants can be informed about their recruitment arm and study results only 1 year after randomization.

### Ethics and dissemination

The study will be conducted following the Good Clinical Practices of the International Conference on Harmonization Good Clinical Practices (ICH-GCP) as well as local and national regulations. The results of the study will be published in scientific journals and presented at scientific meetings. Personal information about enrolled participants will be collected only through REDCap platform.

### Dissemination Policy

Results will be published and be accessible to healthcare professionals and the public.

## Data Availability

This is the trial design for an upcoming study on COVID-19.
The hypothesis is that inhaled Nitric Oxide significantly improves oxygenation in mechanically ventilated patients affected by SARS-CoV-2 infection.

## Authors’ contributions

Authorship for this trial will be given to key personnel involved in trial design, personnel training, recruitment, data collection, statistical plan an data analysis. There are no publication restrictions. CL, BS, HD, LB were responsible for conceptualizing trial design. CL and LB managed patient safety protocol. CL, BS, HD and LB are responsible for recruitment, enrolment and data collection. AB, RDF are responsible for power calculation, statistical plan and data analysis. CL, BS, HD, LB, trained personnel for the clinical trial and built systems for nitric oxide delivery and monitoring. All authors have critically revised the study protocol and approved the final version. All authors agree to be accountable for the accuracy and integrity of all aspects of this trial.

## Competing interests statement

LB salaries are partially supported by NIH/NHLBI 1 K23 HL128882-01A1.

**Table 1.**
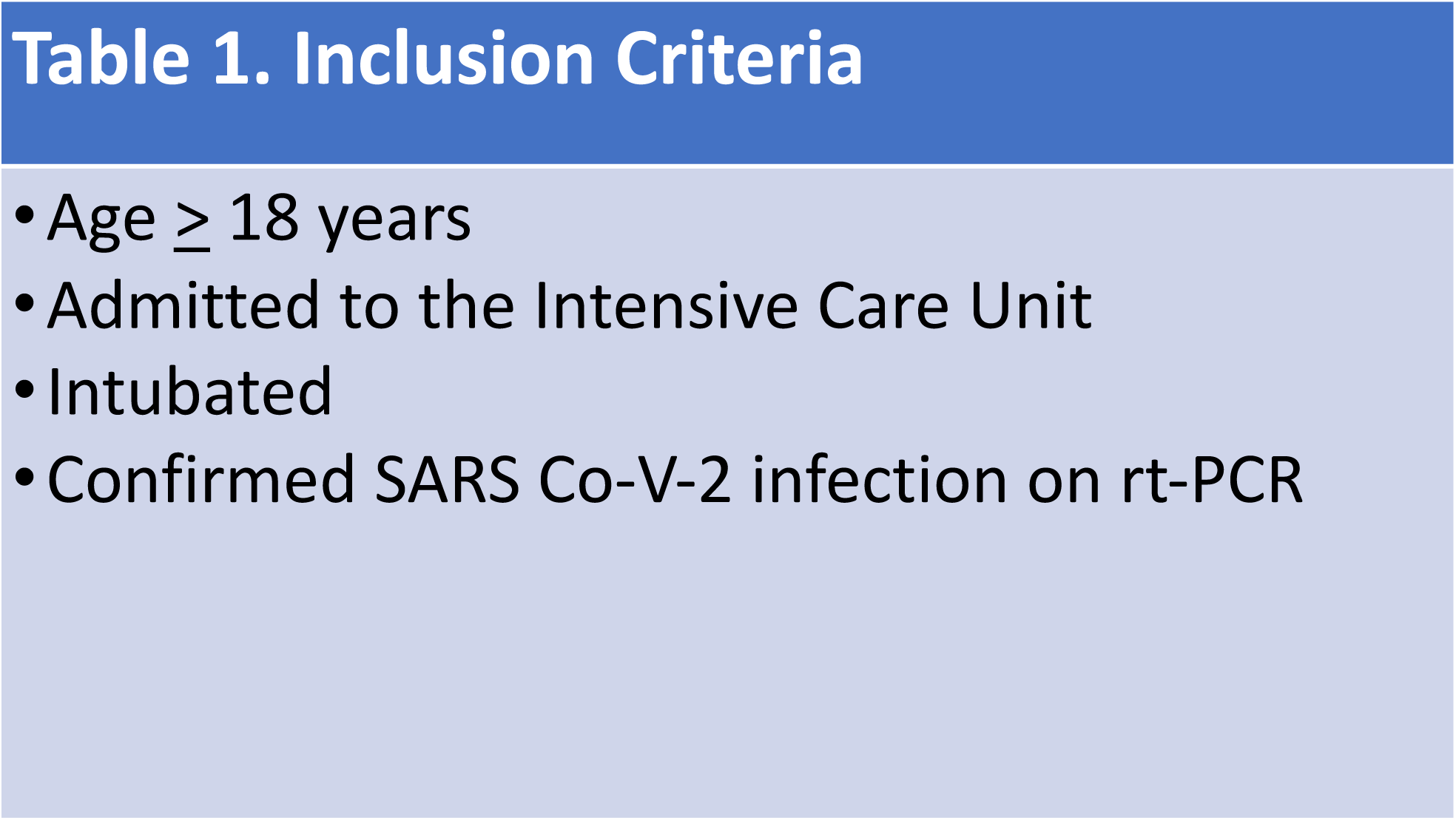
Inclusion Criteria

**Table 2.**
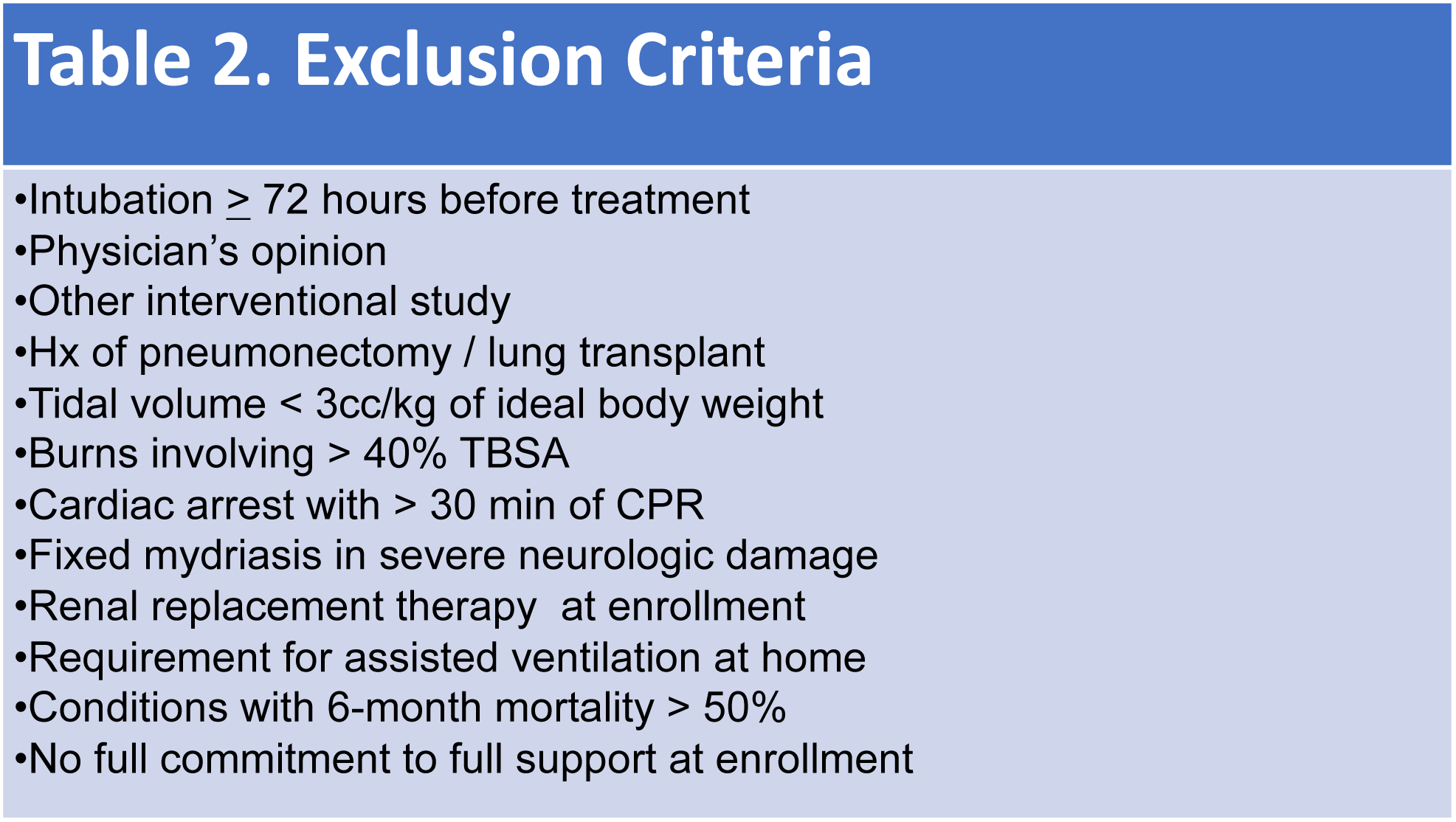
Exclusion Criteria

Flowchart

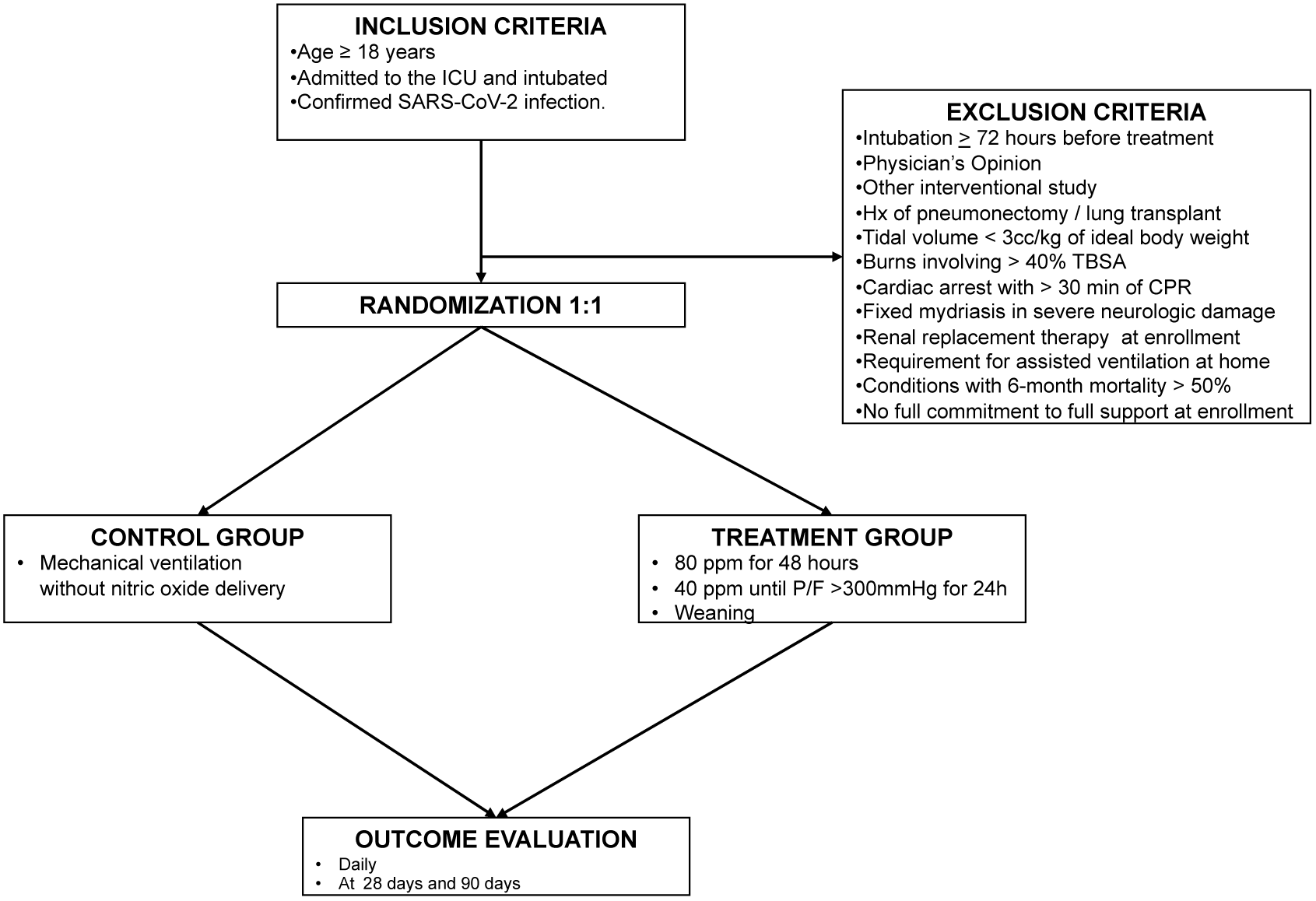

